# Effect of cannabis liberalization on suicide and mental illness following recreational access: a state-level longitudinal analysis in the USA

**DOI:** 10.1101/2020.09.25.20201848

**Authors:** Jacob James Rich, Robert Capodilupo, Michael Schemenaur, Jeffrey A. Singer

## Abstract

**Objective:** To standardize the implementation dates of various cannabis liberalization policies and determine whether previous research by Anderson et al. [D.M. Anderson, D.I. Rees, J.J. Sabia, *American Journal of Public Health* 104, 2369-2376] on medical marijuana access and population-level suicidality is robust to additional years of data and further cannabis liberalization in the form of recreational marijuana access.

**Design:** A state-level longitudinal (panel) analysis. Suicide mortality rates from the National Center for Health Statistics and mental health morbidity rates from the National Survey on Drug Use and Health were employed with the procedures outlined by Anderson et al., using weighted ordinary least squares for three different specifications with various combinations of control variables as a sensitivity analysis to test for robustness.

**Setting:** All 50 states and Washington, DC for the period 1990-2020.

**Participants:** USA population.

**Interventions:** Cannabis liberalization policies in the form of recreational and medical access.

**Primary and Secondary Outcome Measures:** State-level population mental health outcomes in the form of suicide mortality among various age groups for males and females defined by the International Classification of Diseases, Ninth and Tenth Revisions; past-month and -year marijuana use, mental illness, serious mental illness, major depression, and suicidal ideation defined by the Substance Abuse and Mental Health Services Administration.

**Results:** Medical marijuana access was associated with a 3.3% reduction (95% CI -5.0% to -1.7%) in suicide rates for males, which was mediated by a 5.4% reduction (95% CI -8.0% to -2.7%) among males in the 30 to 39 age group. No other mental health outcomes were consistently affected by cannabis liberalization.

**Conclusions:** Adverse mental health outcomes do not follow cannabis liberalization at the state level, confirming the findings of Anderson et al. In addition, there is evidence that medical marijuana access reduces suicide rates for young-adult males.

**Strengths and limitations of this study:**

- Cannabis liberalization policies, which vary greatly throughout the literature, are explicitly defined and corrected from previous studies.
- SAMHSA suppresses state-level geographical information for individual-level responses in the NSDUH, so the analysis relied on population averages for a small number of age groups published in the NSDUH State Prevalence Estimates, which did not allow us to evaluate gender differences for mental health outcomes.
- The reliability of suicide and NSDUH data to estimate true population rates is highly debated.
- Population-level analyses of longitudinal data can be evaluated with multiple accepted methods from the medical literature and it is not clear whether weighted ordinary least squares is the most appropriate approach for this type of analysis.

**Funding statement:** This research received no specific grant from any funding agency in the public, commercial, or not-for-profit sectors.

**Competing interests statement:** Reason Foundation is a 501(c)(3) nonprofit organization completely supported by voluntary contributions from individuals, foundations, corporations, and the sale of its publications. Reason Foundation’s general support includes contributions from marijuana manufacturers, which account for less than one percent of its annual budget.

**Data sharing statement:** Most data relevant to the study are publicly available and included as supplementary information. Mortality rates calculated from death counts of less than 10 deaths for any region are suppressed and may require special permissions for access.

## INTRODUCTION

In the USA 19 states and the District of Columbia have regulated marijuana for recreational use, while medical marijuana flower is available in 36 states (appendix). Cannabis liberalization has gained evolving support towards full legalization, with 60% of American adults now supporting recreational legalization and 91% supporting medical legalization. [1] In October 2018, Canada became the first G7 country to federally legalize marijuana for recreational use. [2] That same month Mexico’s Supreme Court ruled marijuana prohibition unconstitutional, compelling the legislature to pass a marijuana legalization bill. [3] Amid unprecedented support for cannabis legalization in North America, the 117^th^ Congress in the USA is considering such legislation, [4] and policymakers are weighing the public health implications of increasing access to a psychoactive substance that could lead to substance use disorder.

As cannabis use has gained popularity in the USA, [5] global health researchers have increasingly investigated potential adverse outcomes. A 2008 analysis of 2033 participants in the Young in Norway longitudinal study concluded “exposure to marijuana by itself does not lead to depression but that it may be associated with later suicidal thoughts and attempts.” [6] This study joined 13 other longitudinal studies in a 2014 systematic review, which found depressive disorders may be associated with heavy marijuana use. [7] A 2015 study using community-based samples from the Australian Twin Registry found a modest association of marijuana use with suicidal thoughts and attempts, concluding the association between suicidal thoughts and behavior with marijuana use disorder requires further study. [8] A 2017 retrospective study of discordant twins found suicidal thoughts and major depressive disorders were associated with marijuana use and not solely attributable to predisposing factors, [9] confirming similar findings from ten years earlier. [10] In 2020, an evaluation of the National Health and Nutrition Examination Survey found that people reporting depression tend to use marijuana at higher rates. [11]

However, other reports dispute negative mental health outcomes from cannabis liberalization. In particular, a 2014 regression analysis of state-level data from the National Vital Statistics System (NVSS) Mortality Detail Files from 1990-2007 by Anderson et al. found no general association between medical marijuana access and suicide rates, but found 10.8% and 9.4% reductions in the suicide rates in males age 20 to 29 and 30 to 39 in the USA, respectively. [12] Later studies employing difference-in-differences and synthetic control methods on similar data found modest negative associations between medical marijuana access and suicide in young men. [13] [14] In addition, a 2018 evaluation of 465 male and 444 female patients from the Genetics of Opioid Addiction (GENOA) and Determinants of Suicidal Behavior: Conventional and Emergent Risk (DISCOVER) studies found no correlation between suicidal behavior and heavy marijuana use in male or female patients with psychiatric disorders. [15] With the mental health outcomes from marijuana use contested by much of the literature, policymakers would benefit from research investigating the population health effects following increased access to recreational marijuana.

Marijuana use and mental health status are tracked and published by the Substance Abuse and Mental Health Services Administration (SAMHSA). In 2020, about 49.6 million people (or 17.9 percent of the population above the age of 12) consumed marijuana at least once that year in the USA—a historic high. Of those marijuana users, 2.5 million initiated marijuana use for the first time. [5] Marijuana use has been increasing every year since Colorado and Washington voted to legalize marijuana in 2012, a year in which 12.9 percent of the national population reported marijuana use during the past year. [16] Marijuana use in the USA, after reaching an all-time low in 1992, trended slightly upward until 2007, and has steadily increased at a higher rate through 2020. [17] [18]

Reports of major depressive episodes have increased since 2010, while serious thoughts of suicide and severe mental illness have increased each year since those questions were standardized in the survey beginning in 2009. [5] In addition, suicide rates in the USA have steadily increased from an all-time low in 1999 towards an all-time high in 2018. [19] In 2019, the suicide rate in the USA dropped for the first time in 20 years and dropped 5.6% further in 2020 despite the COVID-19 pandemic. [20] With population-level mental health outcomes deteriorating during a period of increasing regular cannabis use, the American Medical Association continues to oppose cannabis legalization for both medical and recreational purposes. [21] But with increasing evidence of marijuana’s medicinal benefits and its growing popularity as a recreational drug, it is necessary to definitively answer whether adverse population health outcomes predictively follow liberalization efforts.

The advent of recreational marijuana laws in the years since Anderson et al. reviewed data from 1990-2007 has provided unprecedented access to cannabis. Previous research has attempted to measure the consequences of new cannabis liberalization efforts, [22] [23] [24] [25] [26] [27] [28] [29] but there are major disparities in the policy implementation dates among these studies. Recent research has confirmed that suicidal behaviors and other adverse mental health outcomes are correlated with marijuana use disorder (MUD), [30] [31] which can be exacerbated by increasing access to cannabis, [32] but these correlations are also prevalent with other substance use disorders. [18] With marijuana use disorder rates stable since 2002, it is not clear that increasing rates of cannabis consumption through liberalization efforts are leading to adverse population-level mental health outcomes. [5] Significant gaps remain in the knowledge of the potential harms and benefits of cannabis use, and the effects these laws have on various facets of mental illness are a fertile topic for new research. [33]

## METHODS

### Settings, participants, procedures

We focused our analysis on state-level mortality data from the National Center for Health Statistics (NCHS) for the period 1990 through 2020 and mental health morbidity rates from the National Survey on Drug Use and Health (NSDUH) for various subsets of the total period based on the duration of each question. All medical certifiers (physicians, coroners, and medical examiners) in the USA reported cause-of-death as defined by the International Classification of Disease to the NCHS, with the Ninth Revision (ICD-9) defining data from 1990-1998 and the Tenth Revision (ICD-10) defining data from 1999-2020. Public deidentified data were retrieved from the National Bureau of Economic Research (NBER) and Wide-ranging ONline Data for Epidemiologic Research (WONDER) databases. [19] [34] For the mental illness data, SAMHSA interviews a target sample size of 67,500 population-representative individuals annually with various questions to estimate rates of drug use and mental health morbidities, which are deidentified to be published as individual responses with no geographical identifiers and as variously defined aggregate rates at the state level.

### Measures

#### Suicide mortality and mental illness morbidity

Our main outcome of interest was the per capita state-level suicide mortality rate (ICD-9: E950-E959; ICD-10: X60-X84, Y87.0, and U03) per 100000 people among the total population, males, females, and both sexes separately for the age groups 15-19, 20-29, 30-39, 40-49, 50-59, and 60 and above. The secondary outcomes of interest were past-month and -year marijuana use and mental health morbidity, which we defined as the percentages of the population that reported “serious thoughts of suicide in the past year”, “any mental illness in the past year”, and “serious mental illness in the past year” for the age groups 18-25, 18 and up, and 26 and up (2009 through 2020); and the percentages of the population that reported they “had at least one major depressive episode in the past year” for the age groups 12-17, 18-25, 18 and up, and 26 and up (2006 through 2020). All cohorts are dynamic, with individuals exiting and entering various age groups each year.

#### Policy variables

Our main exposure of interest takes the form of a cannabis liberalization vector in the form of two indicator variables for recreational and medical marijuana access at the state level. We also control for marijuana decriminalization, 0.08 blood alcohol content (BAC), and zero-tolerance youth drunk driving laws in the covariates. Because there are major discrepancies for when marijuana and drunk driving policies went into effect in the medical literature, [35] we reviewed and contrasted the dates from previous studies and government resources, which we detail in the appendix. Incorrect dates were often used in earlier studies and are now endemic throughout the marijuana and drunk driving literatures. For example, multiple studies list Maryland as becoming a medical marijuana access state as early as October 2003, [24] [32] [36] [37] [38] [39] [40] [41] [42] when the “Darrell Putman Compassionate Use Act” removed the possibility of a jail sentence for marijuana possession if the defendant had a doctor’s note, but did not otherwise protect medical marijuana patients from criminal prosecution. Additionally, some studies previously listed Alabama or Louisiana as medical marijuana states, [42] [43] [44] [45] when “Leni’s Law” in Alabama only decriminalized cannabidiol with low levels of tetrahydrocannabinol (THC) and Louisiana did not allow access to dry marijuana flower until January 1, 2022.

Due to variations in regulations that cause large disparities in the proliferation of medicinal and recreational dispensaries across states, we define the effective indicator date as the day when a typical adult reasonably complying with the new state law would not expect to be arrested for possessing marijuana for personal consumption. Additionally, we only evaluate medical marijuana laws that allow the possession of “smokable” dry cannabis flower that contains THC and can cause euphoric effects.

Marijuana decriminalization laws are defined as the absence of jail time for a first-time offense for possessing up to approximately an ounce of dried cannabis, while recreational marijuana access laws are defined as the protection from all penalties for approximately the same amount of marijuana. For decriminalization and recreational access, quantifying these definitions has been clear since Oregon became the first state to decriminalize marijuana in 1973. In *State v. Twilleager*, [46] the defendant of a marijuana charge in Oregon conceded he had committed the crime of cultivating marijuana on July 21, 1973, prior to the October 5, 1973 effective date of a “decriminalization” amendment reducing the punishment for the first-time offense of such a crime to a $100 fine. He argued that his 6-month jail sentence was “excessive and beyond the jurisdiction of the Court”, since his sentencing took place on January 3, 1974, after the amendment’s effective date. His appeal was struck down after the court cited *Bradley v. United States*, [47] where the Supreme Court affirmed a minimum punishment of 5 years in prison for the conviction of a federal drug charge sentenced on May 6, 1971, when the law was amended to remove such a minimum sentence on May 1, 1971. The case set a precedent that marijuana users should not expect relaxed sentencing from any liberalization policy unless they are charged after the new law’s official effective date.

Medical marijuana access often requires implementing a registry, and laws can take effect long before any state resident can expect protection from previous penalties. For most states, we define a law’s effective date as the day when residents can apply to join the registry, since the laws often explicitly state that one must be registered in order to avoid prosecution for marijuana possession. But this approach does not apply in all jurisdictions, especially in states that regulated medical marijuana in earlier years and did not make protection contingent on registration, instead requiring a simple doctor’s note. However, some more-recent laws allowed for the possession of medical marijuana before any database was established, such as Massachusetts, where Chapter 369 of the 2012 Acts reads “Until the approval of final regulations, written certification by a physician shall constitute a registration card for a qualifying patient.” States also sometimes allowed for those who possessed registration cards from other states to possess marijuana before implementing their own databases, and if this option was available to state residents, we defined the indicator date as the day that policy was effective. This contrasts with states like South Dakota, which only allowed residents from other states with medical marijuana cards to possess cannabis while it was establishing its own marijuana patient registration system.

Because federal law supersedes state law in the USA, marijuana is technically illegal in the entire United States and federal law enforcement may prosecute many marijuana-related crimes regardless of location, such as simultaneous firearm and marijuana possession. However, liberalization efforts have directed many state and local officers to tolerate state-regulated marijuana industries that are federally illegal. The indicator variables take the value of 1 for every full year of enactment and for the first year assume a value between 0 and 1 equal to the proportion of days the policies were effective. [48] Since medical marijuana access and decriminalization are lesser forms of full recreational marijuana access, those indicator variable values equal the recreational marijuana values if they were not previously enacted.

#### Demographic covariates

In harmony with Anderson et al., we control for observable state-level characteristics the literature considers risk factors for suicide: marijuana decriminalization laws, unemployment rates, income per capita, beer excise taxes, 0.08 BAC drunk driving laws, and zero-tolerance drunk driving laws for drivers under the age of 21. Also following Anderson et al., we attempt to control for unobserved factors with indicator variables for time effects, fixed effects, and state-specific linear time trends.

Unemployment rates calculated by the Census Bureau for the Bureau of Labor Statistics (BLS) are retrieved from the IPUMS CPS database, [49] income per capita in 2020 dollars from the Bureau of Economic Analysis (BEA), [50] beer excise tax figures from the Tax Policy Center at the Urban Institute and Brookings Institution, [51] and drunk driving laws from various articles that cite the National Highway Traffic Safety Administration (NHTSA). [48] [52] [53] (appendix)

### Statistical Analysis

Employing the same techniques as Anderson et al., we used population-weighted ordinary least squares (OLS) regressions to conduct a state-level longitudinal (panel) analysis for various subsets of up to 1581 state-years to estimate the effect a recreational and medical marijuana access vector had on the outcome groups. To test the robustness of our findings, we followed Anderson et al. and employed a sensitivity analysis involving three separate specifications for each dependent variable: first only adjusting for state and year effects; then adjusting for state effects, year effects, and covariates; and finally adjusting for state effects, year effects, covariates, and state-specific linear time trends in the most rigorous model. We also corrected standard errors by clustering at the state level for all specifications. Additionally, we followed Anderson et al. by taking the natural log transform of the dependent variables. Because the dependent variables were logged, the estimates can be transformed into percentages by exponentiating, subtracting 1, and multiplying by 100. Since the log of zero is undefined, we dropped all observations of zero suicides. If the coefficients for the marijuana law indicators remain directionally stable and significant for all regression specifications at the *P* < .05 level with confidence intervals that do not include zero, there is evidence that a predictive relationship exists between cannabis liberalization and the mental health outcome variables, and we report the transformed estimates from the most rigorous specification. [54, 55]

## RESULTS

Table 1 presents the dates that various marijuana laws went into effect as of the publication of this paper. Due to delays that are common among states moving to liberalize marijuana, we didn’t include dates for states that have scheduled to liberalize marijuana in the future, like Alabama. Figures 1 and 2 use the recreational marijuana access law dates to plot the rate of reported marijuana use in the past-month and -year over the year of law change, graphing marijuana use for ages 12 to 17 and 18 and up for the nine jurisdictions that had provided recreational marijuana access for at least three years by 2020 (Alaska, California, Colorado, District of Columbia, Maine, Massachusetts, Nevada, Oregon, and Washington). Marijuana use among adults tended to increase following recreational access, but marijuana use among teens remained relatively stable.

**Table 1.** Effective dates of marijuana liberalization

| State | Recreational Marijuana | Medical Marijuana | Decriminalized Marijuana | BAC 0.08 | Zero Tolerance |
| --- | --- | --- | --- | --- | --- |
| Alabama |  |  |  | 1995-10-01 | 1996-05-28 |
| Alaska | 2015-02-24 | 1999-03-04 | *See note | 2001-09-01 | 1996-08-10 |
| Arizona | 2020-11-30 | 2011-04-14 |  | 2001-08-31 | 1990-06-28 |
| Arkansas |  | 2017-06-30 |  | 2001-08-13 | 1993-08-12 |
| California | 2016-11-09 | 1996-11-06 | 1976-01-01 | 1990-01-01 | 1994-01-01 |
| Colorado | 2012-12-10 | 2001-06-01 | 1975-07-01 | 2004-07-01 | 1997-07-01 |
| Connecticut | 2021-07-01 | 2012-10-01 | 2011-07-01 | 2002-07-01 | 1995-10-01 |
| Delaware |  | 2011-07-01 | 2016-01-18 | 2004-07-12 | 1995-07-01 |
| District of Columbia | 2015-02-26 | 2013-07-29 | 2014-07-17 | 1999-04-13 | 1994-05-24 |
| Florida |  | 2019-03-18 |  | 1994-01-01 | 1997-01-01 |
| Georgia |  |  |  | 2001-07-01 | 1997-07-21 |
| Hawaii |  | 2000-06-14 | 2020-01-11 | 1995-06-30 | 1997-12-01 |
| Idaho |  |  |  | 1997-07-01 | 1994-07-01 |
| Illinois | 2020-01-01 | 2014-09-02 | 2016-07-29 | 1997-07-02 | 1995-01-01 |
| Indiana |  |  |  | 2001-07-01 | 1997-01-01 |
| Iowa |  |  |  | 2003-07-01 | 1995-07-01 |
| Kansas |  |  |  | 1993-07-01 | 1997-01-01 |
| Kentucky |  |  |  | 2000-10-01 | 1996-10-01 |
| Louisiana |  | 2022-01-01 | 2021-08-01 | 2003-09-30 | 1997-07-15 |
| Maine | 2017-01-30 | 1999-12-23 | 1976-04-01 | 1988-08-04 | 1995-09-29 |
| Maryland |  | 2017-04-17 | 2014-10-01 | 2001-09-30 | 1990-05-29 |
| Massachusetts | 2016-12-15 | 2013-01-01 | 2009-01-02 | 1994-06-27 | 1994-06-27 |
| Michigan | 2018-12-06 | 2008-12-04 |  | 2003-09-30 | 1994-11-01 |
| Minnesota |  | 2022-03-01 | 1976-04-11 | 2005-09-01 | 1993-06-01 |
| Mississippi |  | 2022-06-01 | 1977-07-01 | 2002-07-01 | 1998-07-01 |
| Missouri |  | 2019-06-28 | 2017-01-01 | 2001-09-29 | 1996-08-28 |
| Montana | 2021-01-01 | 2004-11-02 |  | 2003-04-15 | 1995-10-01 |
| Nebraska |  |  | 1979-01-01 | 2001-09-01 | 1994-01-01 |
| Nevada | 2017-01-01 | 2001-10-01 | 2001-10-01 | 2003-09-23 | 1997-07-16 |
| New Hampshire |  | 2015-12-28 | 2017-09-16 | 1994-01-01 | 1993-01-01 |
| New Jersey | 2021-02-22 | 2012-08-09 |  | 2004-01-20 | 1992-12-17 |
| New Mexico | 2021-06-29 | 2007-07-01 | 2019-07-01 | 1994-01-01 | 1994-01-01 |
| New York | 2021-03-31 | 2015-12-23 | 1977-07-29 | 2003-07-01 | 1996-11-01 |
| North Carolina |  |  | 1977-07-01 | 1993-10-01 | 1995-09-15 |
| North Dakota |  | 2018-10-29 | 2019-08-01 | 2003-08-01 | 1997-07-01 |
| Ohio |  | 2016-09-08 | 1976-07-01 | 2003-06-30 | 1994-05-04 |
| Oklahoma |  | 2018-08-25 |  | 2001-07-01 | 1996-11-01 |
| Oregon | 2015-07-01 | 1998-12-03 | 1973-10-05 | 1983-10-15 | 1991-07-01 |
| Pennsylvania |  | 2017-11-01 |  | 2003-09-30 | 1996-08-02 |
| Rhode Island |  | 2006-01-03 | 2013-04-01 | 2000-07-13 | 1995-06-30 |
| South Carolina |  |  |  | 2003-08-19 | 1993-07-01 |
| South Dakota |  | 2021-11-08 |  | 2002-07-01 | 1998-07-01 |
| Tennessee |  |  |  | 2003-07-01 | 1993-07-01 |
| Texas |  |  |  | 1999-09-01 | 1997-09-01 |
| Utah |  | 2018-12-03 |  | 1983-08-01 | 1992-07-01 |
| Vermont | 2018-07-01 | 2006-07-01 | 2013-07-01 | 1991-07-01 | 1997-09-01 |
| Virginia | 2021-07-01 | 2021-07-01 | 2020-07-01 | 1994-07-01 | 1994-07-01 |
| Washington | 2012-12-06 | 1998-12-03 |  | 1999-01-01 | 1994-07-01 |
| West Virginia |  | 2020-03-05 |  | 2004-05-05 | 1994-06-12 |
| Wisconsin |  |  |  | 2003-09-30 | 1997-10-14 |
| Wyoming |  |  |  | 2002-07-01 | 1998-07-01 |

\*1975-09-02 to 1991-02-04. On September 2, 1975 a decriminalization bill went into effect in Alaska. However, *Ravin v. State* was ruled on May 27, 1975, which effectively legalized marijuana use in one's home before the bill took effect. On February 4, 1991, Alaska recriminalized marijuana through a ballot initiative named Measure 2.

**Figure 1.**
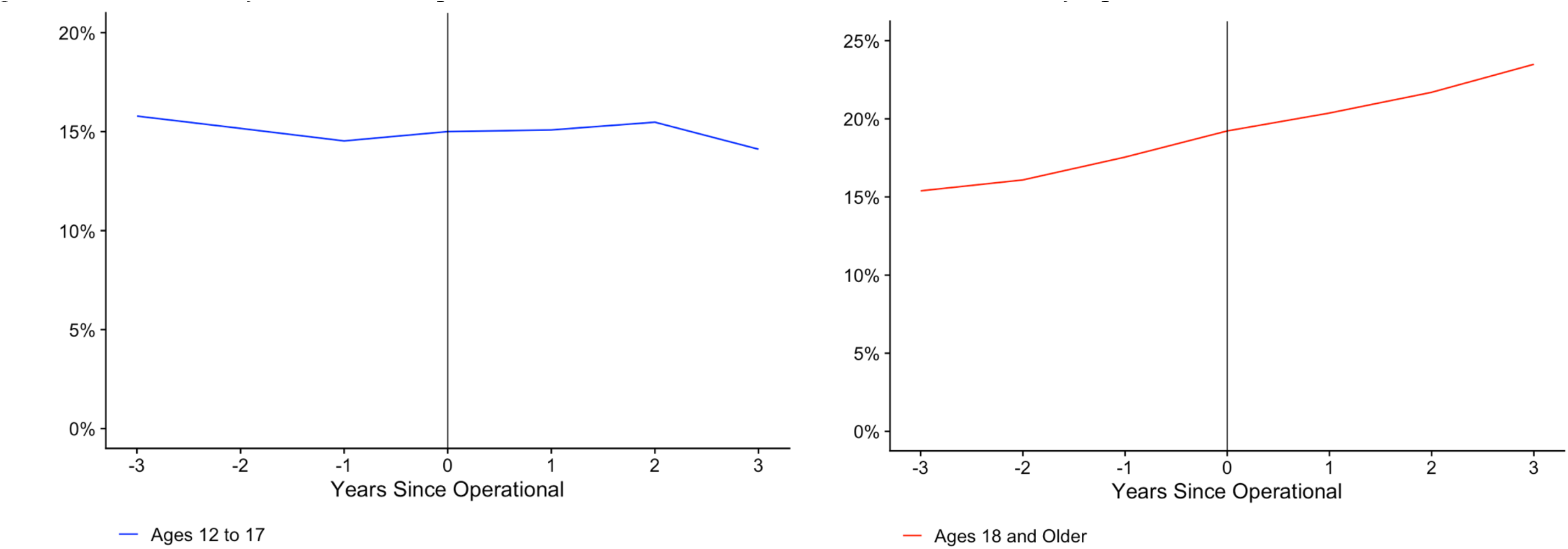
Past-Year Marijuana Use Among 9 Jurisdictions Before and After Recreational Access by Age

**Figure 2.**
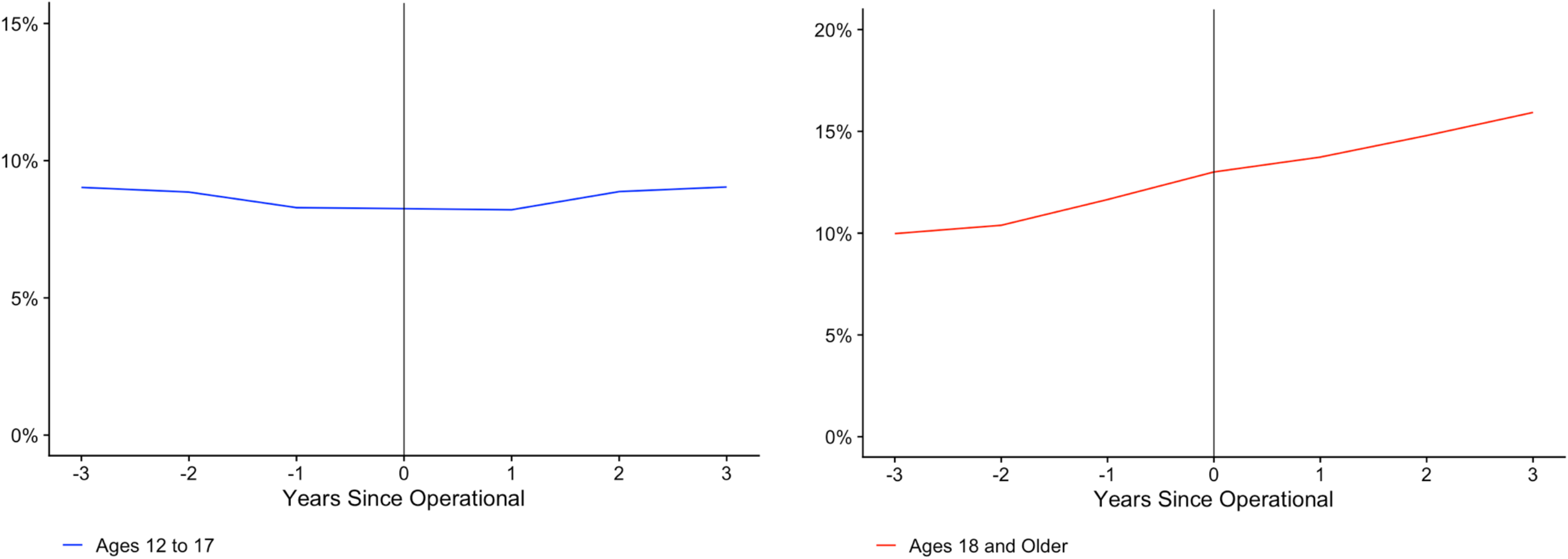
Past-Month Marijuana Use Among 9 Jurisdictions Before and After Recreational Access by Age

Tables 2 and 3 present the main regression results measuring the effect marijuana liberalization had on the mental health outcomes, without transforming the estimates into percentages. The only outcomes that passed each of the three robustness check specifications were a 3.3% reduction (95% CI -5.0% to -1.7%) in suicide rates for males and a 5.4% reduction (95% CI -8.0% to -2.7%) among males in the 30 to 39 age group following medical marijuana access. No other mental health outcomes were consistently affected by cannabis liberalization.

**Table 2.** Suicide Regression Estimates from Anderson et al. (1990-2007), the Replication (1990-2007) and Extension (1990-2020)

| Population | Marijuana Policy | Controls, TE, FE, ST<br>Coefficient (95% CI) |  |  | Controls, TE, FE<br>Coefficient (95% CI) |  |  | TE, FE<br>Coefficient (95% CI) |  |  |
| --- | --- | --- | --- | --- | --- | --- | --- | --- | --- | --- |
|  |  | Anderson et al. | Replication | Extension | Anderson et al. | Replication | Extension | Anderson et al. | Replication | Extension |
| All | Recreational |  |  | -0.012 (-0.031, 0.0061) |  |  | -0.025 (-0.052, 0.0006) |  |  | -0.056** (-0.094, -0.019) |
|  | Medical | -0.049 (-0.099, 0.001) | -0.047 (-0.097, 0.0011) | -0.028** (-0.046, -0.010) | -0.072* (-0.143, -0.0002) | -0.066*** (-0.098, -0.033) | -0.018 (-0.039, 0.0017) | -0.084 (-0.183, 0.016) | -0.081*** (-0.118, -0.045) | -0.024 (-0.048, 0.0004) |
| Male | Recreational |  |  | -0.016 (-0.035, 0.0033) |  |  | -0.019 (-0.043, 0.0038) |  |  | -0.049** (-0.081, -0.016) |
|  | Medical | -0.047* (-0.089, -0.005) | -0.045* (-0.087, -0.002) | -0.034*** (-0.051, -0.017) | -0.065* (-0.128, -0.002) | -0.058*** (-0.087, -0.028) | -0.021* (-0.040, -0.002) | -0.073 (-0.159, 0.014) | -0.070*** (-0.104, -0.037) | -0.026* (-0.048, -0.004) |
| Female | Recreational |  |  | -0.001 (-0.039, 0.0354) |  |  | -0.040 (-0.086, 0.0056) |  |  | -0.080** (-0.141, -0.020) |
|  | Medical | -0.060 (-0.144, 0.024) | -0.056 (-0.151, 0.0388) | -0.000 (-0.035, 0.0335) | -0.080 (-0.180, 0.021) | -0.079* (-0.141, -0.017) | -0.005 (-0.037, 0.0272) | -0.106 (-0.247, 0.036) | -0.106*** (-0.167, -0.045) | -0.011 (-0.049, 0.0262) |
| Male Age 15-19 | Recreational |  |  | -0.040 (-0.132, 0.0520) |  |  | 0.0270 (-0.041, 0.0953) |  |  | -0.013 (-0.085, 0.0580) |
|  | Medical | -0.118 (-0.348, 0.111) | -0.124 (-0.280, 0.0312) | -0.070* (-0.129, -0.012) | -0.091 (-0.252, 0.069) | -0.099* (-0.186, -0.011) | -0.044 (-0.091, 0.0029) | -0.077 (-0.232, 0.078) | -0.097* (-0.180, -0.013) | -0.053* (-0.101, -0.006) |
| Male Age 20-29 | Recreational |  |  | 0.0019 (-0.039, 0.0435) |  |  | 0.0409* (0.0045, 0.0774) |  |  | -0.010 (-0.047, 0.0268) |
|  | Medical | -0.114** (-0.188, -0.038) | -0.067 (-0.163, 0.0280) | -0.051** (-0.081, -0.020) | -0.096** (-0.142, -0.050) | -0.083** (-0.132, -0.033) | -0.051*** (-0.079, -0.022) | -0.096** (-0.147, -0.045) | -0.092*** (-0.138, -0.045) | -0.067*** (-0.097, -0.038) |
| Male Age 30-39 | Recreational |  |  | 0.0033 (-0.038, 0.0451) |  |  | -2.257 (-0.036, 0.0365) |  |  | -0.066** (-0.108, -0.024) |
|  | Medical | -0.099* (-0.175, -0.024) | -0.087** (-0.146, -0.028) | -0.055*** (-0.083, -0.027) | -0.129** (-0.193, 0.065) | -0.108*** (-0.149, -0.068) | -0.062*** (-0.090, -0.034) | -0.147** (-0.244, -0.051) | -0.137*** (-0.177, -0.096) | -0.084*** (-0.115, -0.053) |
| Male Age 40-49 | Recreational |  |  | -0.045* (-0.088, -0.001) |  |  | -0.065** (-0.108, -0.021) |  |  | -0.112*** (-0.169, -0.054) |
|  | Medical | -0.060 (-0.149, 0.030) | -0.068 (-0.143, 0.0060) | -0.060*** (-0.092, -0.029) | -0.113* (-0.223, -0.004) | -0.110*** (-0.161, -0.059) | -0.054** (-0.088, -0.019) | -0.129 (-0.297, 0.039) | -0.135*** (-0.196, -0.075) | -0.060** (-0.101, -0.019) |
| Male Age 50-59 | Recreational |  |  | -0.008 (-0.045, 0.0290) |  |  | -0.070** (-0.113, -0.026) |  |  | -0.081** (-0.136, -0.026) |
|  | Medical | -0.015 (-0.099, 0.068) | -0.017 (-0.091, 0.0562) | -0.002 (-0.029, 0.0251) | -0.010 (-0.101, 0.080) | -0.012 (-0.063, 0.0372) | 0.0108 (-0.018, 0.0397) | -0.030 (-0.155, 0.095) | -0.028 (-0.080, 0.0234) | 0.0184 (-0.013, 0.0505) |
| Male Age 60 Up | Recreational |  |  | 0.0075 (-0.027, 0.0427) |  |  | -0.052** (-0.089, -0.015) |  |  | -0.055* (-0.100, -0.011) |
|  | Medical | 0.036 (-0.020, 0.094) | 0.0485* (0.0009, 0.0960) | 0.0047 (-0.019, 0.0294) | -0.010 (-0.079, 0.058) | -0.017 (-0.056, 0.0200) | 0.0136 (-0.009, 0.0367) | -0.008 (-0.082, 0.065) | -0.011 (-0.046, 0.0247) | 0.0187 (-0.005, 0.0429) |
| Female Age 15-19 | Recreational |  |  | -0.052 (-0.206, 0.1010) |  |  | -0.137 (-0.288, 0.0145) |  |  | -0.097 (-0.245, 0.0498) |
|  | Medical | 0.083 (-0.388, 0.554) | -0.114 (-0.368, 0.1401) | -0.101 (-0.207, 0.0051) | -0.105 (-0.403, 0.225) | -0.177* (-0.332, -0.022) | -0.042 (-0.130, 0.0460) | -0.069 (-0.388, 0.249) | -0.145* (-0.277, -0.013) | -0.016 (-0.101, 0.0681) |
| Female Age 20-29 | Recreational |  |  | 0.0174 (-0.072, 0.1072) |  |  | -0.000 (-0.074, 0.0729) |  |  | -0.040 (-0.111, 0.0310) |
|  | Medical | -0.008 (-0.138, 0.122) | -0.026 (-0.161, 0.1083) | -0.003 (-0.060, 0.0532) | -0.044 (-0.174, 0.087) | 0.0064 (-0.078, 0.0916) | 0.0080 (-0.039, 0.0560) | -0.062 (-0.185, 0.060) | -0.038 (-0.114, 0.0385) | -0.004 (-0.052, 0.0422) |
| Female Age 30-39 | Recreational |  |  | 0.0457 (-0.041, 0.1328) |  |  | -0.005 (-0.081, 0.0702) |  |  | -0.071 (-0.144, 0.0014) |
|  | Medical | -0.035 (-0.212, 0.141) | -0.035 (-0.166, 0.0968) | 0.0024 (-0.057, 0.0624) | -0.110* (-0.218, -0.003) | -0.122** (-0.213, -0.031) | -0.015 (-0.065, 0.0335) | -0.125* (-0.243, -0.007) | -0.135** (-0.220, -0.050) | -0.036 (-0.087, 0.0150) |
| Female Age 40-49 | Recreational |  |  | -0.062 (-0.138, 0.0144) |  |  | -0.115** (-0.187, -0.043) |  |  | -0.172*** (-0.266, -0.079) |
|  | Medical | -0.041 (-0.151, 0.069) | -0.025 (-0.146, 0.0966) | 0.0111 (-0.043, 0.0657) | -0.078 (-0.211, 0.056) | -0.068 (-0.153, 0.0164) | -0.029 (-0.079, 0.0200) | -0.120 (-0.332, 0.092) | -0.117** (-0.205, -0.029) | -0.035 (-0.093, 0.0232) |
| Female Age 50-59 | Recreational |  |  | 0.0040 (-0.079, 0.0874) |  |  | -0.056 (-0.145, 0.0316) |  |  | -0.108* (-0.202, -0.014) |
|  | Medical | -0.104 (-0.225, 0.018) | -0.108 (-0.295, 0.0792) | 0.0444 (-0.010, 0.0991) | -0.019 (-0.164, 0.125) | -0.055 (-0.150, 0.0388) | 0.0307 (-0.018, 0.0798) | -0.068 (-0.275, 0.140) | -0.075 (-0.165, 0.0151) | 0.0250 (-0.026, 0.0766) |
| Female Age 60 Up | Recreational |  |  | 0.0019 (-0.061, 0.0655) |  |  | -0.027 (-0.100, 0.0455) |  |  | -0.057 (-0.146, 0.0322) |
|  | Medical | -0.121* (-0.240, -0.004) | -0.131 (-0.274, 0.0107) | -0.034 (-0.083, 0.0153) | -0.082 (-0.199, 0.036) | -0.104* (-0.193, -0.015) | -0.016 (-0.061, 0.0283) | -0.082 (-0.208, 0.043) | -0.096* (-0.174, -0.018) | -0.015 (-0.062, 0.0301) |

\* $P < 0.05$ , \*\* $P < 0.01$ , \*\*\* $P < 0.001$

**Table 3.** NSDUH Mental Health Regression Estimates

| Outcome | Marijuana Policy | Controls. TE. FE. ST | Controls. TE. FE | TE. FE | Period |
| --- | --- | --- | --- | --- | --- |
|  |  | Coefficient (95% CI) | Coefficient (95% CI) | Coefficient (95% CI) |  |
| Depression Age 12 to 17 | Recreational | -0.043** (-0.075, -0.012) | 0.0112 (-0.022, 0.0447) | 0.0152 (-0.018, 0.0492) | 2006-2020 |
|  | Medical | -0.026 (-0.055, 0.0019) | -0.013 (-0.038, 0.0113) | -0.003 (-0.027, 0.0212) |  |
| Depression Age 18 to 25 | Recreational | 0.0263 (-0.027, 0.0804) | 0.0304 (-0.019, 0.0801) | 0.0213 (-0.032, 0.0750) | 2006-2020 |
|  | Medical | -0.001 (-0.033, 0.0312) | 0.0124 (-0.016, 0.0409) | 0.0182 (-0.008, 0.0453) |  |
| Depression Age 26 Up | Recreational | 0.0053 (-0.033, 0.0444) | 0.0516** (0.0174, 0.0858) | 0.0463** (0.0176, 0.0749) | 2006-2020 |
|  | Medical | -0.008 (-0.038, 0.0213) | -0.007 (-0.031, 0.0166) | -0.004 (-0.027, 0.0186) |  |
| Depression Age 18 Up | Recreational | 0.0122 (-0.019, 0.0438) | 0.0476*** (0.0201, 0.0750) | 0.0421*** (0.0188, 0.0654) | 2006-2020 |
|  | Medical | -0.008 (-0.034, 0.0186) | -0.003 (-0.025, 0.0174) | 0.0004 (-0.020, 0.0210) |  |
| Suicidal Thoughts Age 18-25 | Recreational | -0.003 (-0.043, 0.0364) | 0.0160 (-0.028, 0.0609) | -0.020 (-0.059, 0.0184) | 2009-2020 |
|  | Medical | 0.0159 (-0.014, 0.0466) | 0.0106 (-0.017, 0.0386) | 0.0012 (-0.027, 0.0298) |  |
| Suicidal Thoughts Age 26 Up | Recreational | 0.0327 (-0.007, 0.0733) | 0.0563** (0.0151, 0.0974) | 0.0347 (-0.046, 0.0694) | 2009-2020 |
|  | Medical | 0.0465* (0.0085, 0.0845) | -0.012 (-0.044, 0.0201) | -0.021 (-0.054, 0.0115) |  |
| Suicidal Thoughts 18 Up | Recreational | 0.0214 (-0.012, 0.0554) | 0.0417* (0.0089, 0.0746) | 0.0140 (-0.012, 0.0403) | 2009-2020 |
|  | Medical | 0.0371* (0.0078, 0.0664) | -0.006 (-0.032, 0.0195) | -0.015 (-0.042, 0.0110) |  |
| Mental Illness Age 18 to 25 | Recreational | -0.004 (-0.036, 0.0268) | 0.0046 (-0.023, 0.0327) | 0.0003 (-0.026, 0.0274) | 2009-2020 |
|  | Medical | -0.011 (-0.035, 0.0130) | -0.002 (-0.022, 0.0166) | -0.001 (-0.020, 0.0180) |  |
| Mental Illness Age 26 Up | Recreational | 0.0049 (-0.027, 0.0378) | 0.0334* (0.0034, 0.0634) | 0.0376** (0.0142, 0.0609) | 2009-2020 |
|  | Medical | -0.015 (-0.040, 0.0096) | -0.013 (-0.036, 0.0093) | -0.013 (-0.035, 0.0091) |  |
| Mental Illness Age 18 Up | Recreational | 0.0033 (-0.025, 0.0320) | 0.0287* (0.0017, 0.0558) | 0.0313** (0.0111, 0.0516) | 2009-2020 |
|  | Medical | -0.014 (-0.036, 0.0075) | -0.012 (-0.033, 0.0080) | -0.011 (-0.031, 0.0086) |  |
| Serious Mental Illness Age 18-25 | Recreational | 0.0375 (-0.005, 0.0807) | 0.0442 (-0.011, 0.0999) | 0.0106 (-0.032, 0.0539) | 2009-2020 |
|  | Medical | -0.035 (-0.074, 0.0037) | 0.0091 (-0.026, 0.0452) | 0.0067 (-0.027, 0.0406) |  |
| Serious Mental Illness Age 26 Up | Recreational | -0.005 (-0.051, 0.0406) | 0.0249 (-0.009, 0.0594) | 0.0093 (-0.024, 0.0430) | 2009-2020 |
|  | Medical | 0.0384 (-0.007, 0.0845) | 0.0153 (-0.015, 0.0461) | 0.0028 (-0.028, 0.0346) |  |
| Serious Mental Illness Age 18 Up | Recreational | 0.0001 (-0.039, 0.0395) | 0.0266 (-0.005, 0.0584) | 0.0113 (-0.017, 0.0406) | 2009-2020 |
|  | Medical | 0.0230 (-0.014, 0.0603) | 0.0126 (-0.012, 0.0379) | 0.0041 (-0.022, 0.0304) |  |

\* $P < 0.05$ , \*\* $P < 0.01$ , \*\*\* $P < 0.001$

## DISCUSSION

Despite suicide rates reaching unprecedented levels across the country, suicides have decreased for some demographics following marijuana liberalization. Our analysis finds strong evidence that medical marijuana laws reduce suicide rates for men age 30 to 39, which mediates reductions in suicide for the entire male population. Interestingly, Anderson et al. found that medical marijuana legalization led to 10.8% (95% CI -17.1% to -3.7%) and 9.4% (95% CI -16.1% to -2.4%) drops in the suicide rates for males in the 20 to 29 and 30 to 39 age groups. It should also be noted that the male age 20 to 29 group passed the sensitivity analysis for the extended period and failed only one of three regressions in the replication.

Given that the confidence intervals of Anderson et al.’s statistically significant male cohorts overlap the estimates from this analysis, we conclude that the original study’s model is robust to new developments in marijuana policy and additional years of data. Our study also supports recent research by Anderson et al. (2021) observing that marijuana use among teens is not affected by recreational marijuana access laws. [56] Although the reasons for this are not clear, this development may be due to the proliferation of marijuana dispensaries in states that allow recreational access to marijuana, which may reduce the number of black-market drug dealers who are willing to sell psychoactive substances to children who are still experiencing major cognitive developments. However, without public access to state-level NSDUH estimates of marijuana use that distinguish between genders, it is difficult to determine whether changes in marijuana use mediate decreasing rates of suicide among males and females.

Moreover, we find that marijuana liberalization in general does not lead to undesirable outcomes for any age group or gender at the population-level, despite correlations between marijuana use disorder (MUD) and mental illness at the individual-level. [30] Nonetheless, there is no evidence that the liberalization of marijuana is leading to higher rates of suicide or mental illness in the USA.

### Limitations

SAMHSA suppresses state-level geographical information for individual-level responses in the NSDUH because of privacy concerns. Due to the limitations of public NSDUH data, it is not possible to utilize geographical controls for individual responses to compare our population-level results to potential individual-level analyses. In addition, our mental illness analysis relied on population averages for a small number of age groups published in the NSDUH State Prevalence Estimates, [57] which prevented us from evaluating either gender or racial disparities. This was particularly restricting in this paper, because we reported decreasing average rates of suicide among young adult males following medical marijuana access, but we could not determine whether this was correlated with changes in marijuana use among this cohort. To improve on future public health research independent of the federal government’s efforts, SAMHSA should add state-level identifiers to its public individual-response data for all years of the NSDUH and its predecessor, the National Household Survey on Drug Abuse (NHSDA). Since the surveys’ state-level samples consist of at least hundreds of responses each, privacy will not be a concern if state-of-residence information is released to the public for each individual response. Permitting state-level identifiers would allow future researchers to employ various analytical methods that use geographical identifiers to determine state-level policy changes in individual-level analyses.

There are also many challenges to the accuracy of the NSDUH and suicide rates. Critics claim that because the NSDUH requires the surveyed individuals to have a residence, much of the population are not represented by the estimates, especially those suffering from homelessness. [58] [59] In addition, it is widely believed that suicide rates are underreported. [60] These are legitimate criticisms, but the regression analyses in this study are much more concerned with the changes in rates, as opposed to the systematic inaccuracies of the rates themselves that are overly consistent over time. Since there is no reason to believe that states with recreational or medical marijuana access would be more or less likely to misreport suicides or mental illness, and consistent methods for collecting these data were employed for the entire period of interest for all dependent variables, we are confident that the coefficients reflect the relationships between evolving trends of cannabis availability and the mental health identifiers of interest.

However, population-level analyses of longitudinal (panel) data can be evaluated with multiple accepted methods from the medical literature. It is not clear that OLS is the most appropriate approach for this type of analysis and there’s no objective method of determining which approach is preferable. Prominent similarly-designed analyses employing OLS have been replicated and extended, only to show that the results were not robust to additional years of data. For example, the observation that medical marijuana laws reduce opioid overdoses reversed after seven additional years of data were added to the regressions. [43] The replicating authors described the observations as spurious, since only 2.5% of the study population used medical cannabis, which they argued would unlikely contribute to significant changes in opioid-related mortality in either direction. Our analysis also replicated the approach of a highly cited study, using thirteen additional years of data, but instead confirmed the original results.

## CONCLUSIONS

The use of any drug entails certain risks along with any benefits. Cannabis is no exception. Critics of marijuana legalization point to studies showing correlations between heavy cannabis use and suicide, depression, and mental health disorders. [30] However, such studies demonstrating correlation have yet to confirm causation, which should be determined by a model’s ability to predict. [61] Although those reporting depression to SAMHSA have increasingly used marijuana since states began increasing access to regulated cannabis, [11] we observe no evidence that cannabis liberalization has predictive relationships with reports of any mental illness. In addition, medical marijuana access is consistently followed by reductions in suicide for some male age groups. Anderson et al. found similar relationships between medical marijuana access and suicide rates. The lack of an association between mental health and recreational marijuana access in our study is likely because those who would have benefited the most from marijuana consumption likely had the highest demand for cannabis, making them more likely to benefit from medical access relative to those who started consuming marijuana following recreational access. In this analysis, the recreational access indicators measure the marginal gain from moving from medical access to recreational access, meaning that the majority of the effect of recreational marijuana is absorbed by the medical indicator and the coefficient for recreational marijuana is an underestimate of its total effect. [62]

We propose as medicinal and recreational use of marijuana becomes more widespread and mainstream, concerns about the correlation between marijuana use and depression should not interfere with state or federal efforts to decriminalize or legalize cannabis. In fact, legalization will have the salutary effect of allowing more rigorous research—now inhibited by federal prohibition—into the further benefits, as well as any other potential harms, from the long-term use of marijuana, and promote safer use.

## Supporting information

Supplemental Data and Policy Sources

## Data Availability

Most data relevant to the study are publicly available and included as supplementary information. Mortality rates calculated from death counts of less than 10 deaths for any region are suppressed and may require special permissions for access.

## Contributors

JJR designed the study, collected and organized the data, and wrote the majority of the paper. RC collected data for comparison purposes and provided useful suggestions. MS reviewed individual-level SAMHSA data and relayed invaluable descriptive statistics to describe marijuana users. JAS conducted a literature review, wrote much of the introduction, and supervised the research.

